# Impact of surgical approach on Stage I Small Cell Lung Cancer according to the Eighth Edition of TNM Classification: A Population-Based Study

**DOI:** 10.1101/2025.03.24.25324388

**Authors:** Bin Wang, Jian Sun, Hui Zhu, Hongbo Guo, Xiaokang Guo

## Abstract

**Background:** Limited evidence exists regarding the comparative effectiveness of lobectomy (L) and sublobar resection (sub-L) for early-stage small cell lung cancer (SCLC).

**Methods:** We identified patients with stage I (pT1-T2aN0M0) SCLC who underwent definitive surgery from the Surveillance, Epidemiology, and End Results (SEER) database between 2004 and 2019. Overall survival (OS) and lung cancer-specific death (LCSD) were assessed using Kaplan-Meier analysis, Cox regression model analysis, and competing risk model analysis. Propensity score matching (PSM) was employed to mitigate bias.

**Outcomes:** Among the 560 patients in this study, 383 underwent lobectomy, 177 patients underwent sublobar resection, with 138 undergoing wedge resections (WR) and 39 undergoing segmental resection (SR). L demonstrated better OS than sub-L for resected stage I SCLC before and after PSM (5-year OS: 52.2% vs. 37.8%, p < 0.001; 54.3% vs. 37.9%, p=0.025). LCSD did not significantly differ between L and sub-L groups before and after PSM (5-year LCSD: 36.5% vs. 43.0%, p=0.088; 34.5% vs. 39.1%, p=0.444). Multivariate Cox regression analysis (hazard ratio: 0.65, 95% CI 0.5 - 0.85, p = 0.002) and competing risk regression model analysis (sub-distribution hazard ratio: 0.76, 95% CI 0.57 – 1.02, P=0.07) further confirmed above conclusion. In subgroup analysis, L was associated with improved OS and reduced LCSD compared to sub-L among patients with pure SCLC.

**Conclusion:** Lung cancer-specific survival was similar between L and sub-L groups in early-stage SCLC. Sub-L was deemed suitable for specifically selected patients with SCLC.

## 1. Introduction

SCLC is characterized by rapid growth, high malignancy, and a propensity for mediastinal lymph node and distant metastasis, distinguishing it from non-small-cell lung cancer (NSCLC) [1]. SCLC constitutes approximately 15% of all lung cancers [2]. Early randomized controlled trials in 1973 and 1994 found no survival benefit from surgery in SCLC [3, 4]. Chemotherapy in combination with radiation has become the standard treatment for limited SCLC. Recent retrospective studies, based on extensive databases, have indicated the potential benefit of surgery for very early SCLC [5–9]. The National Comprehensive Cancer Network (NCCN) guidelines currently endorse surgery as part of the comprehensive treatment for patients with T1-2N0M0 SCLC [10]. Traditionally, lobectomy has been the primary recommendation for non-small cell lung cancer due to its superior prognosis compared to sublobar resection [11–14]. With the widespread application of chest CT and lung cancer screening, more and more early-stage lung cancers are being detected, leading more and more scholars to question whether sublobar resection, which has the advantages of preserving more lung parenchyma, less postoperative morbidity, and faster recovery, should be the intentional resection method rather than only compromise surgical method for early NSCLC. Japanese researchers recently reported the results of a large randomized trial (JCOG0802), showing that in clinical stage IA patients with tumors ≤2cm, segmental resection is superior to lobectomy in overall survival and not inferior to lobectomy in recurrence-free survival [15]. A subsequent multicenter, international large randomized trial (CALGB140503) similarly showed that sublobar resection was not inferior to lobectomy in recurrence-free survival and overall survival [16]. Based on these two findings, sublobar resection is increasingly used in early NSCLC with tumors ≤2cm. However, there is a lack of prospective studies and limited observational data comparing lobar and sublobar resection in early-stage SCLC [17–20]. The inconsistency in previous research results raises doubts about the reliability of the conclusions. Hence, our study aims to determine the optimal surgical approach for stage I (T1-2aN0M0) SCLC patients by comparing the prognosis of lobectomy and sublobar resection based on the SEER database.

## 2. Material and methods

### 2.1 Data sources

This retrospective study was based on data from the Surveillance, Epidemiology, and End Results (SEER) database pertaining to information on patients diagnosed with cancer between 2004 and 2019 (Incidence-SEER Research Plus Data, 17 Registries, Nov 2021 Sub, 2000-2019). The SEER database did not require ethical approval from the authors’ organizations because it provides free data download services for researchers and ensures confidentiality of the personal information of patients. All methods were performed in accordance with the relevant guidelines and regulations.

### 2.2 Study Population

Clinicopathological and prognostic data was extracted from SEER database, including patients’ age, gender and race; tumor’s size, location, stage and therapeutic details. The inclusion criteria included: one primary SCLC only, diagnosed between 2004 and 2019 from SEER, site recode ICD-O-3/WHO 2008 of “lung and bronchus”, histologic/behavior type codes 8041/3–8045/3), stage I SCLC with tumors ≤4 cm and underwent surgery. The TNM stage of selected patients was reclassified based on the American Joint Commission on Cancer, 8th Edition. Patients were ruled out based on the following criteria: unidentified surgery status, unknown survival time, cases without a positive pathological diagnosis, TNM stage unable to be reclassified, and diagnosis based only on the autopsy/death certificate. Finally, 560 patients were included in the study.

### 2.3 Statistical Analysis

Baseline demographics and clinical characteristics were compared using Pearson χ2 test for discrete variables. A Kaplan-Meier survival curve with the log-rank test were performed for overall survival (OS). Multivariate Cox model was used to compute hazard ratios (HR) and 95% confidence intervals (95% CI) for independent prognostic factors. Fine and Gray’s test was applied to test the difference between the two groups using a cumulative incidence function curve. Additionally, sub-distribution hazard ratio (sHR) and 95% CI were calculated in competing risk models and sensitivity analysis. Propensity score matching (PSM) was used to control for confounding factors affecting lung cancer survival. PSM is a balancing technique to approximate the balance in measured covariates between L and sub-L groups. Propensity scores were calculated using multiple logistic regression analysis with the following covariates: age, gender, race, laterality, primary site, year of diagnosis, pathological subtype, chemotherapy, radiotherapy, dissected regional lymph nodes and TNM stage. The patients were matched in a 1:1 ratio using the caliper method with a matching tolerance of 0.05.

All analyses were achieved by utilizing SPSS software (version 26.0), and R statistical software (version 4.1.2). All statistical tests were two-sided, with P < 0.05 considered to be indicative of statistical significance.

## 3. Results

### 3.1 Patient characteristics

Between 2014 and 2019, we identified 560 patients diagnosed with stage I SCLC and underwent definitive surgery from the SEER database. Of these, sublobar resection was to 117 patients (20.89%), while 383 patients (79.11%) underwent lobectomy. After PSM, 260 patients were evenly distributed into the lobectomy group (n = 130) and the sublobar resection group (n = 130). The baseline characteristics of patients before and after PSM are outlined in Table 1. Significant differences in age, regional lymph node dissection, and adjuvant radiotherapy were observed between the two groups before PSM. Patients who underwent sublobar resection were more likely to be over 65 years old, undergo lymph node resection, and receive postoperative adjuvant radiotherapy (p<0.001). However, after PSM, basic demographics and clinical characteristics were well-balanced between the two groups, as illustrated in Table 2.

**Table 1.**
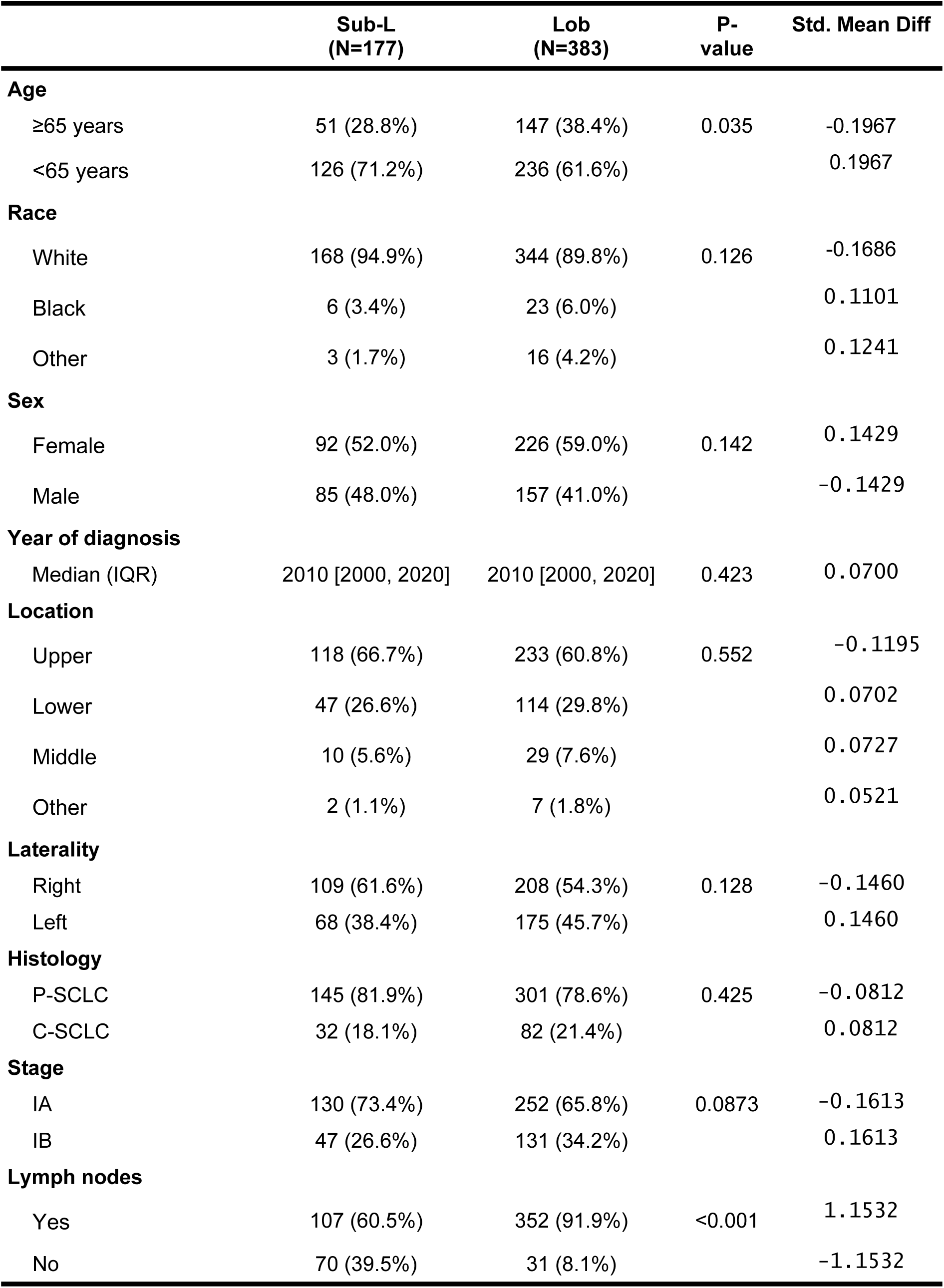
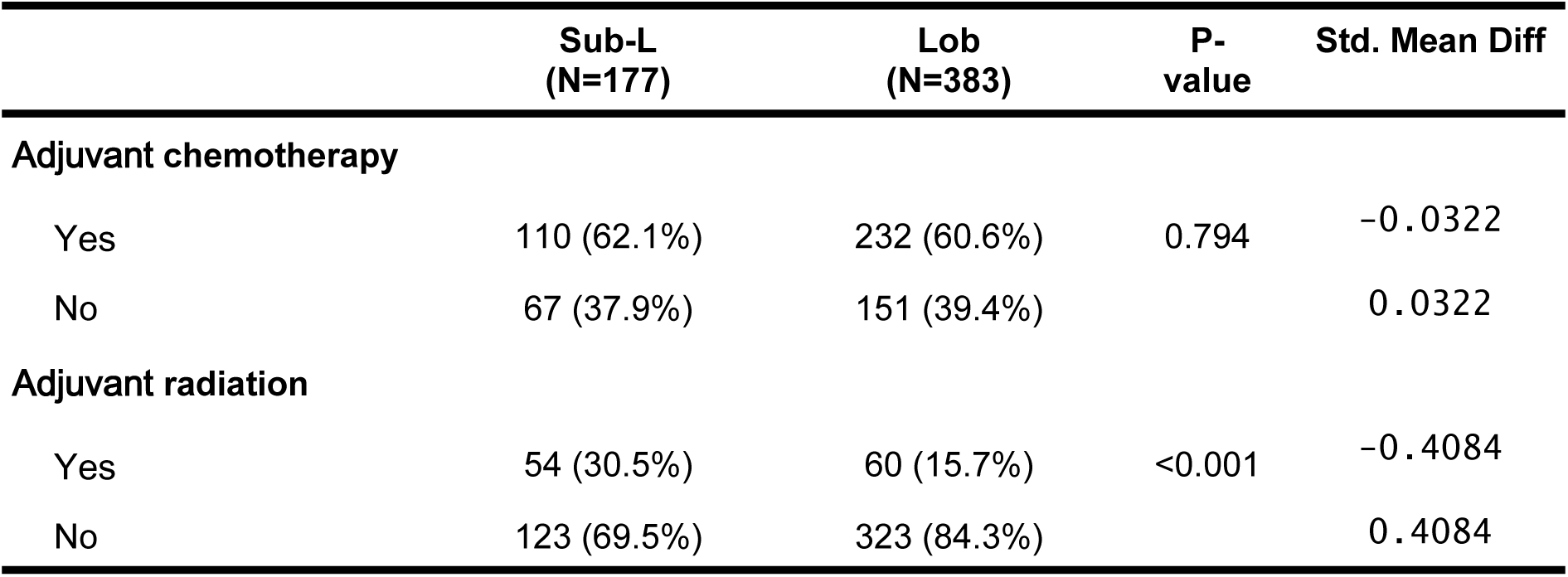
Clinicopathological characteristics of SCLCs before a PSM.

**Table 2.** Clinicopathological characteristics of SCLCs after PSM.

|  | Sub-L<br>(N=130) | Lob<br>(N=130) | P-value | Std. Mean Diff |
| --- | --- | --- | --- | --- |
| Age |  |  |  |  |
| ≥65 years | 43 (33.1%) | 43 (33.1%) | 1 | -0.0158 |
| <65 years | 87 (66.9%) | 87 (66.9%) |  | 0.0158 |
| Race |  |  |  |  |
| White | 123 (94.6%) | 119 (91.5%) | 0.598 | 0.0763 |
| Black | 4 (3.1%) | 7 (5.4%) |  | -0.1295 |
| Other | 3 (2.3%) | 4 (3.1%) |  | 0.0384 |
| Sex |  |  |  |  |
| Female | 66 (50.8%) | 69 (53.1%) | 0.804 | 0.0156 |
| Male | 64 (49.2%) | 61 (46.9%) |  | -0.0156 |
| Year of diagnosis |  |  |  |  |
| Median (IQR) | 2010 [2000, 2020] | 2010 [2000, 2020] | 0.437 | -0.0287 |
| Location |  |  |  |  |
| Upper | 85 (65.4%) | 82 (63.1%) | 0.898 | -0.1734 |
| Lower | 37 (28.5%) | 41 (31.5%) |  | 0.1346 |
| Middle | 6 (4.6%) | 6 (4.6%) |  | 0.0872 |
| Other | 2 (1.5%) | 1 (0.8%) |  | 0.0000 |
| Laterality |  |  |  |  |
| Right | 76 (58.5%) | 76 (58.5%) | 1 | -0.0927 |
| Left | 54 (41.5%) | 54 (41.5%) |  | 0.0927 |
| **Histology** |  |  |  |  |
| P-SCLC | 106 (81.5%) | 106 (81.5%) | 1 | 0.0188 |
| C-SCLC | 24 (18.5%) | 24 (18.5%) |  | -0.0188 |
| **Stage** |  |  |  |  |
| IA | 93 (71.5%) | 84 (64.6%) | 0.287 | 0.0000 |
| IB | 37 (28.5%) | 46 (35.4%) |  | 0.0000 |
| **Lymph nodes** |  |  |  |  |
| Yes | 104 (80.0%) | 105 (80.8%) | 1 | 0.0000 |
| No | 26 (20.0%) | 25 (19.2%) |  | 0.0000 |
| **Adjuvant chemotherapy** |  |  |  |  |
| Yes | 77 (59.2%) | 71 (54.6%) | 0.531 | -0.1417 |
| No | 53 (40.8%) | 59 (45.4%) |  | 0.1417 |
| **Adjuvant radiation** |  |  |  |  |
| Yes | 33 (25.4%) | 35 (26.9%) | 0.888 | -0.0635 |
| No | 97 (74.6%) | 95 (73.1%) |  | 0.0635 |

### 3.2 OS analysis

Patients who underwent lobectomy exhibited significantly longer OS compared to those who underwent sublobar resection before PSM (5-year OS: 52.2% vs. 37.8%, p < 0.001; Figure 1a). This significant difference persisted between the lobectomy and sublobar resection groups after PSM (5-year OS: 54.3% vs. 37.9%, p = 0.025; Figure 1b). In the univariate analysis, not only surgery but also age, gender, year of diagnosis, and regional lymph node dissection were identified as factors significantly associated with OS outcomes. Subsequently, factors with p<0.1 or clinical significance from the univariate analysis were included in the multivariate Cox regression model. In the multivariate analysis, lobectomy was still associated with improved OS (HR 0.65, 95% CI 0.5 - 0.85, p = 0.002) (Table 3).

**Figure 1a.**
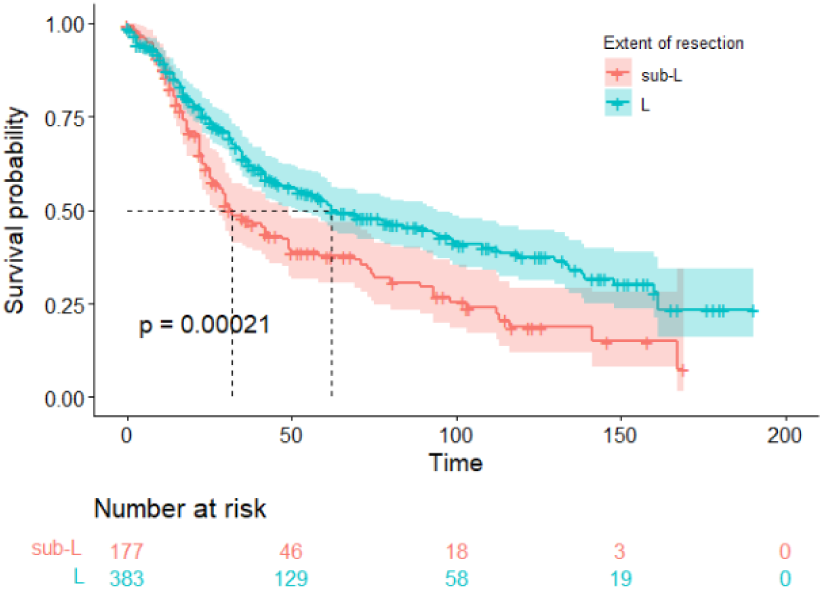
Kaplan–Meier survival curve of stage I SCLC patients before PSM

**Figure 1b.**
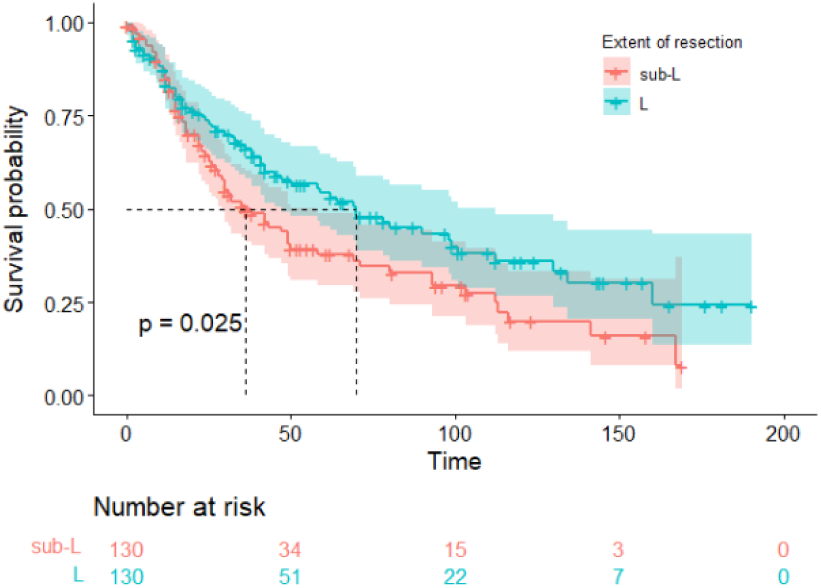
Kaplan–Meier survival curve of stage I SCLC patients after PSM

**Table 3.** Univariate and multivariate Cox regression analyses with OS.

|  | Univariate |  | Multivariate |  |
| --- | --- | --- | --- | --- |
| Characteristics | HR (95% CI) | p | HR (95% CI) | p |
| **Age** |  |  |  |  |
| <65 years | Reference |  | Reference |  |
| ≥65 years | 1.57 ( 1.22 - 2.02 ) | <0.001 | 1.48 (1.14 - 1.92) | 0.003 |
| **Race** |  |  |  |  |
| White | Reference |  | Reference |  |
| Black | 0.54 (0.28-1.04) | 0.065 | 0.58 (0.29 - 1.14) | 0.113 |
| Other | 1.32 (0.70-2.48) | 0.392 | 1.43 (0.75 - 2.72) | 0.276 |
| **Sex** |  |  |  |  |
| Female | Reference |  | Reference |  |
| Male | 1.28 (1.01-1.61) | 0.037 | 1.26 (0.99 - 1.59) | 0.056 |
| **Year of diagnosis** | 0.96 (0.93-0.99) | 0.006 | 0.96 (0.94 - 0.99) | 0.011 |
| **Location** |  |  |  |  |
| Upper | Reference |  |  |  |
| Lower | 1.05 (0.82-1.36) | 0.683 |  |  |
| Middle | 0.65 (0.38-1.12) | 0.124 |  |  |
| Other | 1.18 (0.48-2.87) | 0.717 |  |  |
| **Laterality** |  |  |  |  |
| Right | Reference |  |  |  |
| Left | 1.02 (0.81-1.29) | 0.848 |  |  |
| **Histology** |  |  |  |  |
| P-SCLC | Reference |  |  |  |
| C-SCLC | 1.02 (0.77-1.35) | 0.883 |  |  |
| **Stage** |  |  |  |  |
| IA | Reference |  | Reference |  |
| IB | 1.16 (0.91-1.49) | 0.223 | 1.28 (1 - 1.65) | 0.053 |
| **Lymph nodes** |  |  |  |  |
| Yes | Reference |  | Reference |  |
| No | 1.35 (1.02-1.79) | 0.035 | 1.1 (0.81 - 1.5) | 0.568 |
| **Adjuvant chemotherapy** |  |  |  |  |
| Yes | Reference |  | Reference |  |
| No | 1.24 (0.99-1.57) | 0.066 | 1.09 ( 0.85 - 1.41 ) | 0.499 |
| **Adjuvant radiation** |  |  |  |  |
| Yes | Reference |  | Reference |  |
| No | 1.33 (0.99-1.78) | 0.060 | 1.26 ( 0.92 - 1.73 ) | 0.157 |
| **Extend of surgery** |  |  |  |  |
| sub-L | Reference |  | Reference |  |
| L | 0.64 (0.5-0.81) | <0.001 | 0.65 (0.5 - 0.85) | 0.002 |

### 3.3 LCSD analysis

A competing risk analysis-based cumulative incidence function curve for LCSD for all patients is shown in Figure 2. The lobectomy group showed a similar LCSD (5-year LCSD: 36.5% vs 43.0%, p=0.088) while significantly lower non-LCSD 5-year non-LCSD: 11.3% vs 19.2%, p=0.015 compared with the sublobar resection group before PSM (Figure 2a). No significant difference of LCSD 5-year: 34.5% vs 39.1%, p=0.444 and non-LCSD 5-year non-LCSD: 11.2% vs 23.1%, p=0.090 was found between the L and sub-L groups after PSM (Figure 2b). Univariate analysis, conducted using a Fine–Gray sub-distribution proportional-hazards model, indicated that lobectomy was associated with similar LCSD (sHR:0.78 (0.58, 1.04), p=0.090) but decreased non-LCSD (sHR: 0.64 (0.42, 0.95), p=0.027) in patients with SCLC. Even after adjusting for variables with p<0.1 or clinical significance from the univariate analysis, the multivariate analysis still found that lobectomy was associated with similar LCSD (sHR: 0.76, 95% CI 0.57 – 1.02, p=0.07) and significantly lower non-LCSD (sHR: 0.65, 95% CI 0.43 – 0.96, p=0.032) compared with sublobar resection (Table 4).

**Figure 2a.**
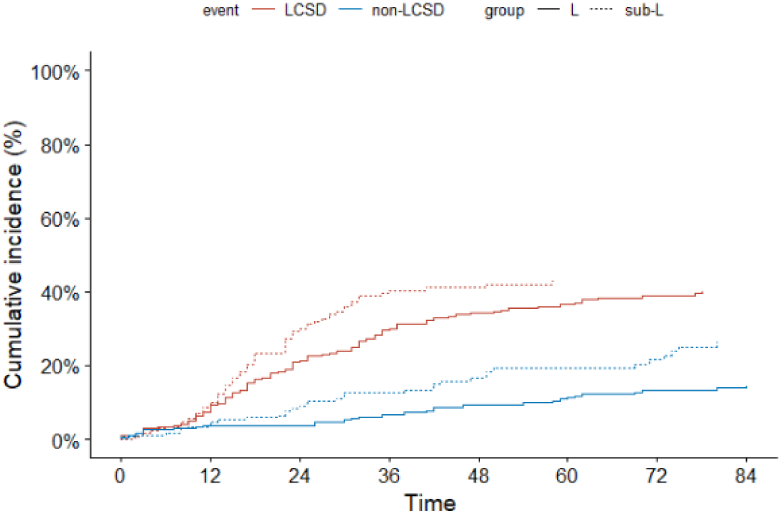
Cumulative incidence curve of stage I SCLC patients before PSM.

**Figure 2b.**
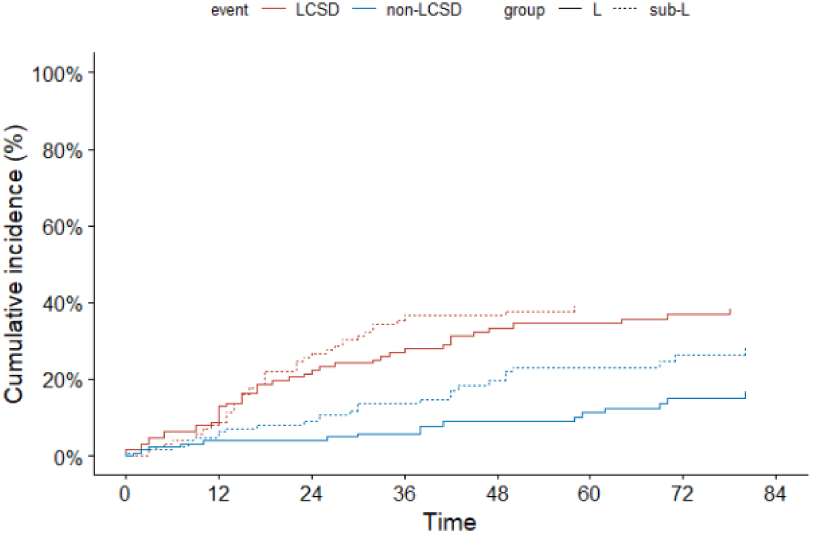
Cumulative incidence function curve of stage I SCLC patients after PSM.

**Table 4.**
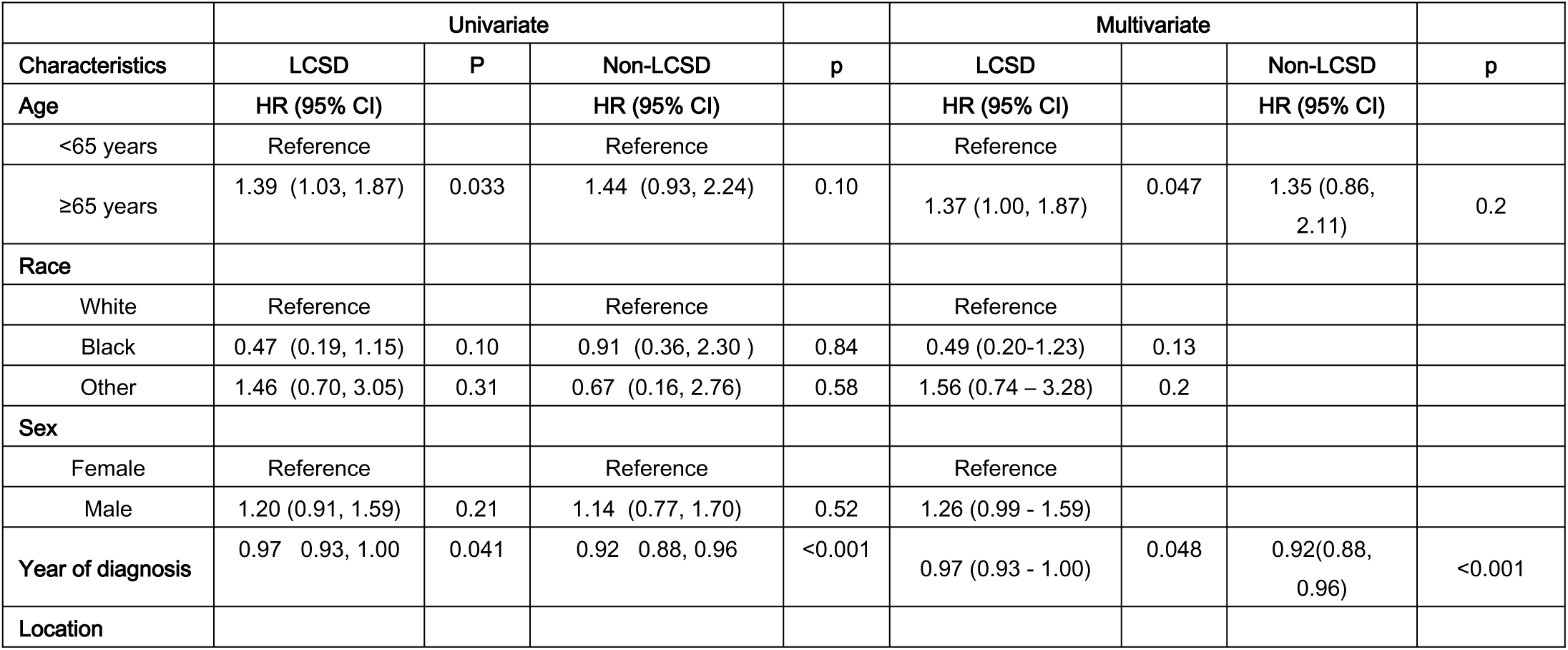
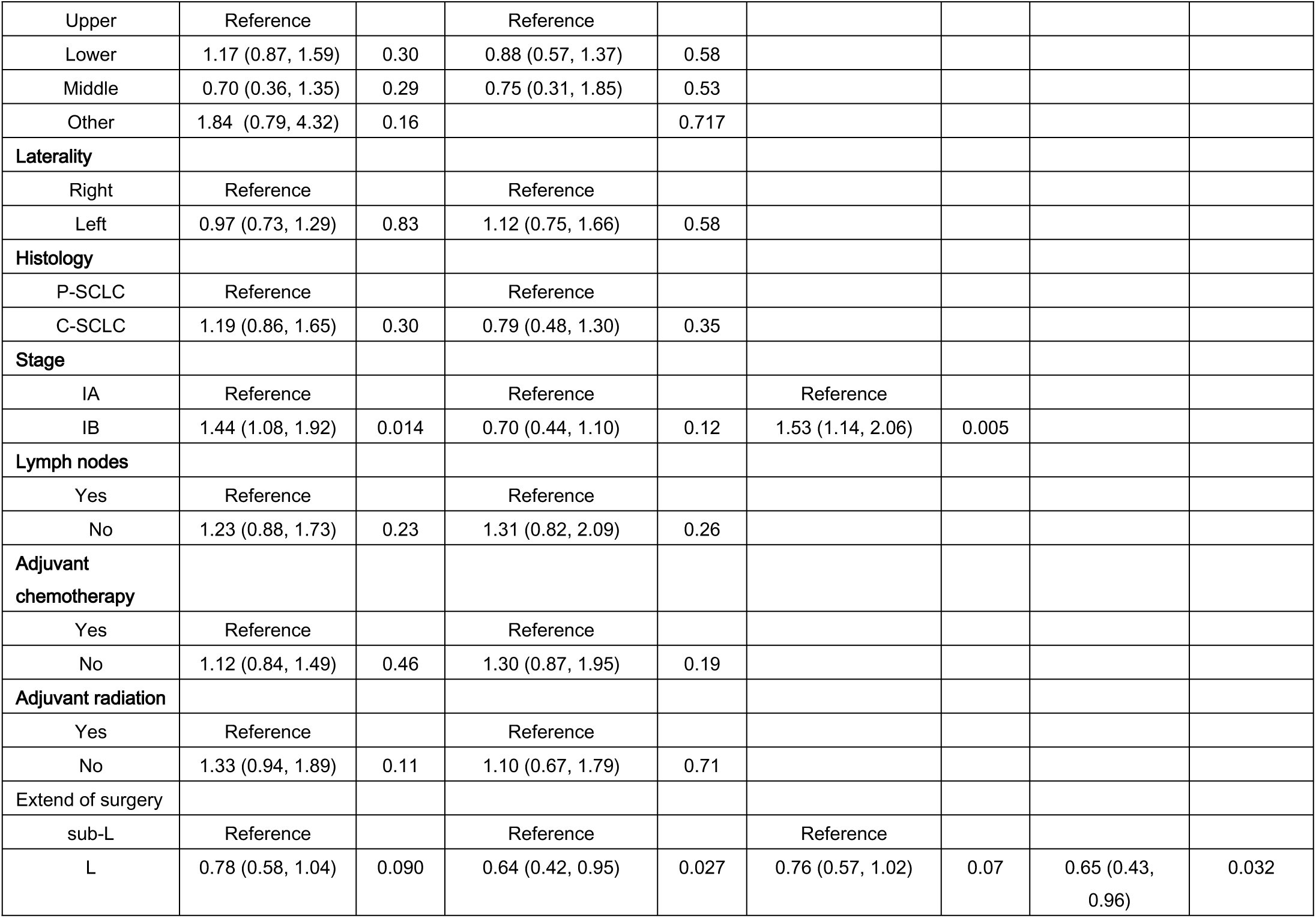
Univariate and multivariate subdistribution proportional hazards model analyses with LCSD.

### 3.4 Analysis among WR, SR and L

Five-year OS was 37.9%, 37.0% and 52% for WR, SR, and L, respectively (p<0.001) (Figure 3a). In multivariable Cox regression, patients undergoing L experienced better survival compared with patients undergoing WR (HR 0.66, 95% CI 0.58 - 0.86, p = 0.005), whereas patients undergoing SR experienced similar survival compared with patients undergoing WR (HR 1.01, 95% CI 0.64 −1.59, p = 0.958). (Table 5)

**Table 5.** Multivariate Cox regression analyses with OS.

|  | Multivariate |  |
| --- | --- | --- |
| Characteristics | HR (95% CI) | p |
| **Age** |  |  |
| <65 years | Reference |  |
| ≥65 years | 1.53 (1.18 - 1.98) | 0.001 |
| **Race** |  |  |
| White | Reference |  |
| Black | 0.95 (0.69 - 1.31) | 0.513 |
| Other | 1.02 (0.83 - 1.72) | 0.767 |
| **Sex** |  |  |
| Female | Reference |  |
| Male | 1.25 (0.99 - 1.58) | 0.060 |
| **Year of diagnosis** | 0.95 (0.92 - 0.97) | 0.013 |
| **Stage** |  |  |
| IA | Reference |  |
| IB | 1.29 (1 - 1.66) | 0.052 |
| **Lymph nodes** |  |  |
| Yes | Reference |  |
| No | 1.13 (0.82 - 1.54) | 0.455 |
| **Adjuvant chemotherapy** |  |  |
| Yes | Reference |  |
| No | 1.08 (0.84 - 1.4) | 0.555 |
| **Adjuvant radiation** |  |  |
| Yes | Reference |  |
| No | 1.28 (0.93 - 1.76) | 0.127 |
| **Extend of surgery** |  |  |
| WR | Reference |  |
| SR | 1.01 (0.64 - 1.59) | 0.958 |
| L | 0.66 (0.49 - 0.89) | 0.006 |

**Figure 3a.**
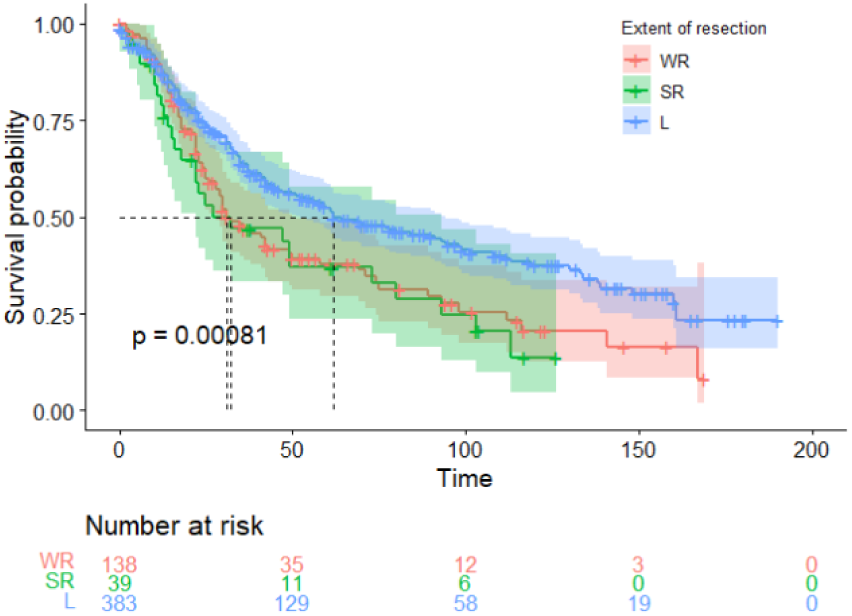
Kaplan–Meier survival curve among WR, SR and L.

Five-year LCSD was 42.0%, 47.6%, and 36.5% for WR, SR, and L, respectively (p=0.080) (Figure 3b). In multivariate analysis, patients undergoing L (sHR: 0.836, 95% CI 0.60 – 1.16, p=0.280) and SR (sHR: 1.26, 95% CI 0.74 – 2.15, p=0.390) both experienced similar survival compared with patients undergoing WR (Table 6)

**Figure 3b.**
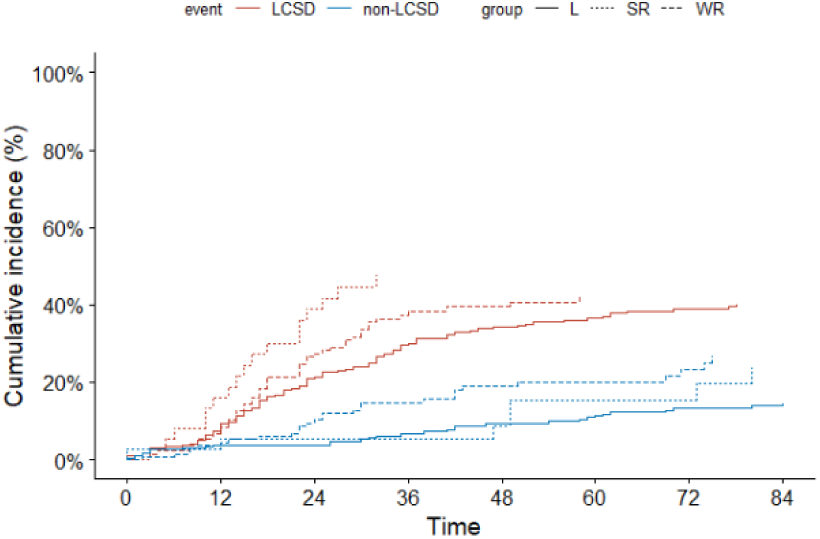
Cumulative incidence function curve among WR, SR and L.

**Table 6.** Multivariate subdistribution proportional hazards model analyses with LCSD.

| Characteristics | Multivariate |  |  | p |
| --- | --- | --- | --- | --- |
|  | LCSD |  | Non-LCSD |  |
| **Age** | **HR (95% CI)** |  | **HR (95% CI)** |  |
| <65 years | Reference |  |  |  |
| ≥65 years | 1.40 (1.04-1.90) | 0.029 | 1.40 (0.89-2.20) | 0.140 |
| **Race** |  |  |  |  |
| White | Reference |  |  |  |
| Black | 0.91 (0.38-2.19) | 0.81 |  |  |
| Other | 0.63 (0.22-2.06) | 0.54 |  |  |
| **Sex** |  |  |  |  |
| Female | Reference |  |  |  |
| Male | 1.15 (0.86 - 1.53) | 0.308 |  |  |
| **Year of diagnosis** |  |  | 0.92(0.88, 0.96) | <0.001 |
| **Stage** |  |  |  |  |
| IA | Reference |  |  |  |
| IB | 1.48 (1.10-1.98) | 0.010 |  |  |
| **Extend of surgery** |  |  |  |  |
| WR | Reference |  |  |  |
| SR | 1.26 (0.74-2.15) | 0.400 | 0.68 (0.30-1.57) | 0.300 |
| L | 0.83 (0.60-1.16) | 0.300 | 0.59 (0.39-0.89) | 0.013 |

### 3.5 Subgroup analysis after PSM

To evaluate the impact of surgical approach on patients at different stages, we categorized individuals into stages IA and IB. Among stage IA patients, lobectomy demonstrated superior OS (5-year OS: 63.7% vs. 44.0%, p = 0.035; Figure 4a) but comparable LCSD (5-year LCSD: 24.9% vs. 35.9%, p = 0.103; Figure 4b) compared to sublobar resection. Conversely, in stage IB patients, lobectomy was associated with similar OS (5-year OS: 39.1% vs. 21.5%, p = 0.130; Figure 4c) and LCSD (5-year LCSD: 50.0% vs. 47.7%, p = 0.790; Figure 4d) to sublobar resection. For survival analyses based on age, patients were stratified into ≥65 years and <65 years. Among patients <65 years, we observed similar OS (5-year OS: 55.7% vs. 49.0%, p = 0.440; Figure 5a) and LCSD (5-year LCSD: 25.3% vs. 30.2%, p = 0.338; Figure 5b) between the L and sub-L resection groups. Among patients ≥65 years, we noted improved OS (5-year OS: 55.6% vs. 32.7%, p = 0.034; Figure 5c) but similar LCSD (5-year LCSD: 39.3% vs. 43.1%, p = 0.836; Figure 5d) in the L group compared to the sub-L group. Moreover, patients were categorized into pure small cell lung cancer (P-SCLC) and combined small cell lung cancer (C-SCLC) based on histological subtype. In P-SCLC, lobectomy was associated with superior OS (5-year OS: 57.8% vs. 31.8%, p = 0.047; Figure 6a) and decreased LCSD (5-year LCSD: 29.1% vs. 45.3%, p = 0.008; Figure 6b) compared to sublobar resection. However, in C-SCLC, lobectomy was linked to worse OS (5-year OS: 39.3% vs. 62.3%, p = 0.047; Figure 6c) and increased LCSD (5-year LCSD: 56.5% vs. 13.3%, p < 0.001; Figure 6d) compared to sublobar resection.

**Figure 4a.**
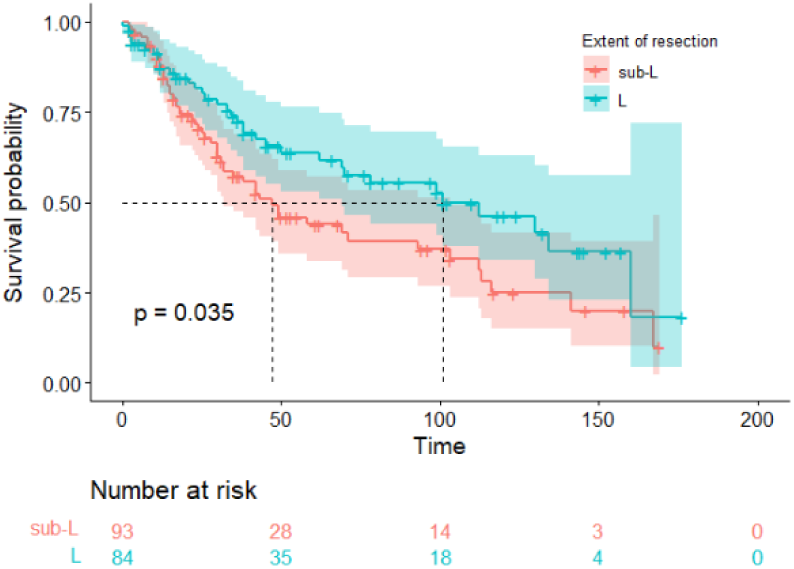
Kaplan–Meier survival curve of stage IA SCLC patients.

**Figure 4b.**
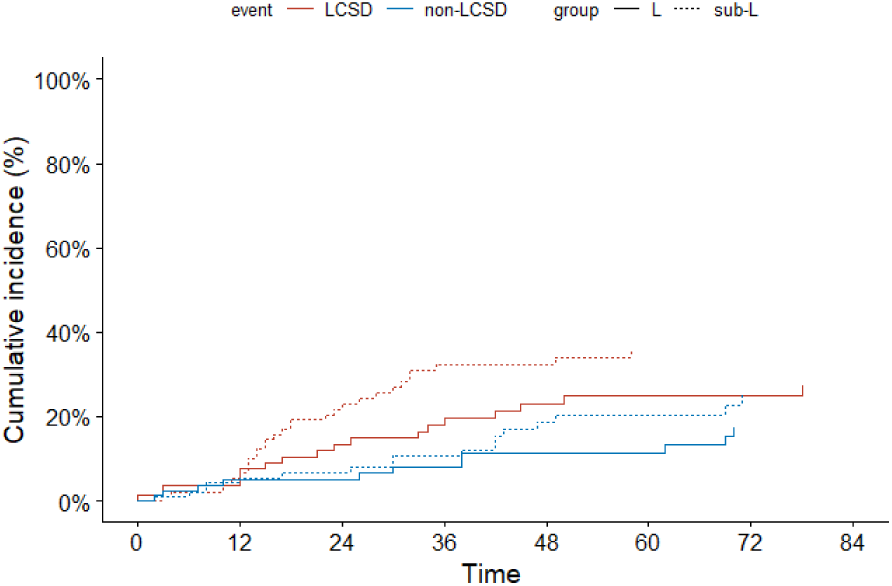
Cumulative incidence function curve of stage IA SCLC patients.

**Figure 4c.**
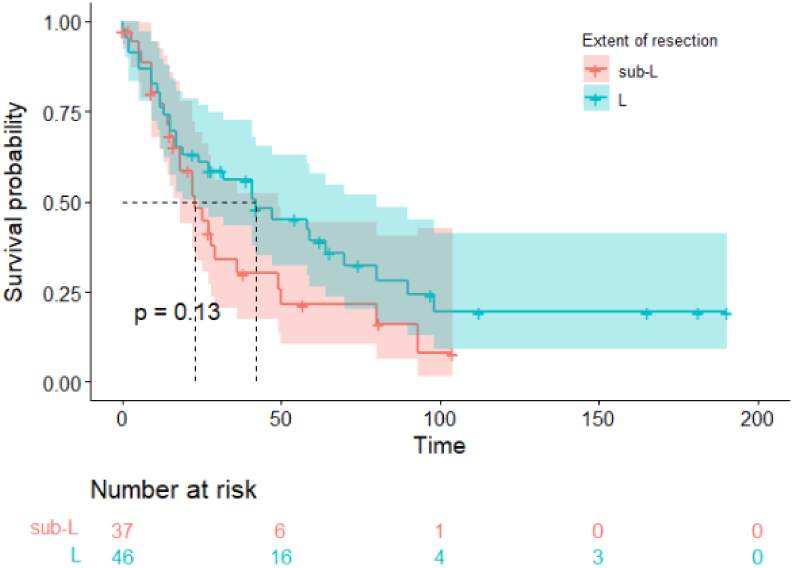
Kaplan–Meier survival curve of stage IB SCLC patients.

**Figure 4d.**
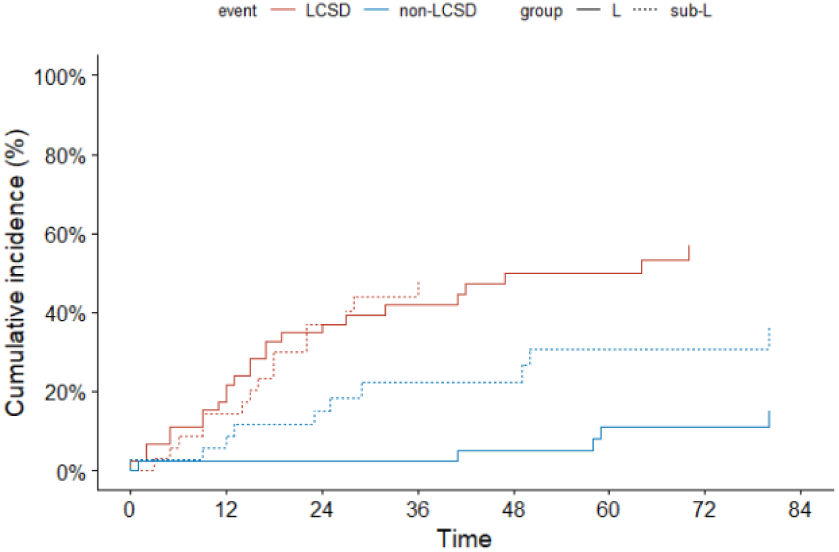
Cumulative incidence function curve of stage IB SCLC patients.

**Figure 5a.**
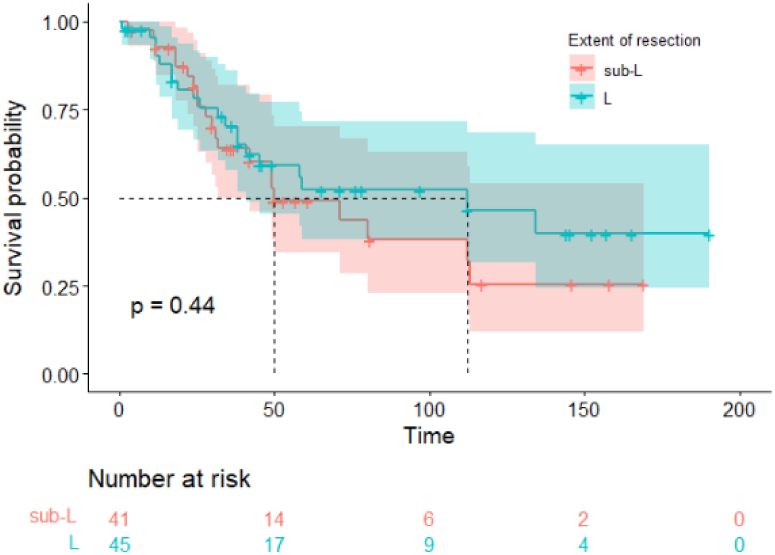
Kaplan–Meier survival curve of stage I SCLC patients with age <65 years.

**Figure 5b.**
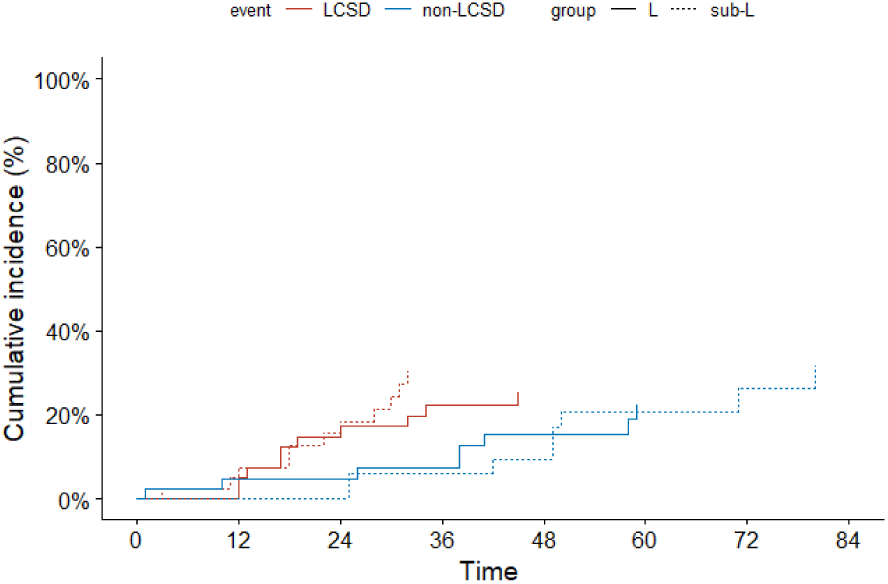
Cumulative incidence function curve of stage I SCLC patients with age <65 years.

**Figure 5c.**
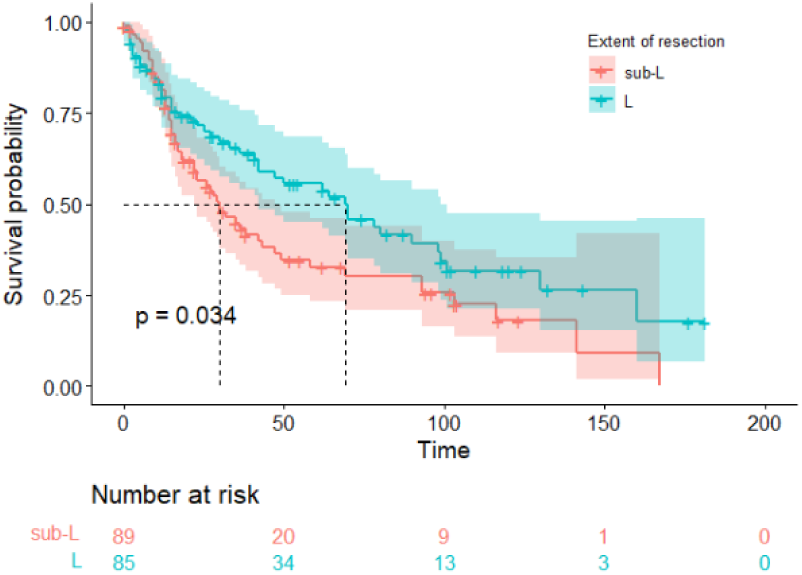
Kaplan–Meier survival curve of stage I SCLC patients with age ≥65 years.

**Figure 5d.**
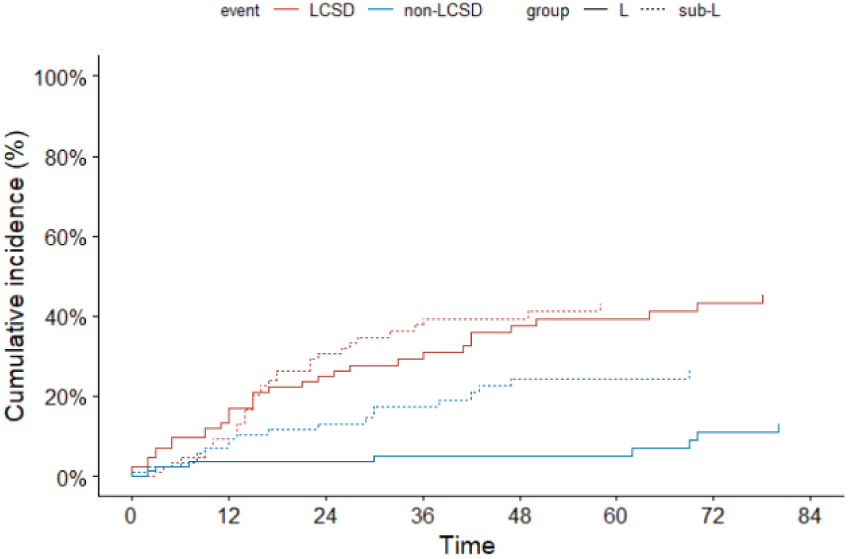
Cumulative incidence function curve of stage I SCLC patients with age ≥65 years.

**Figure 6a.**
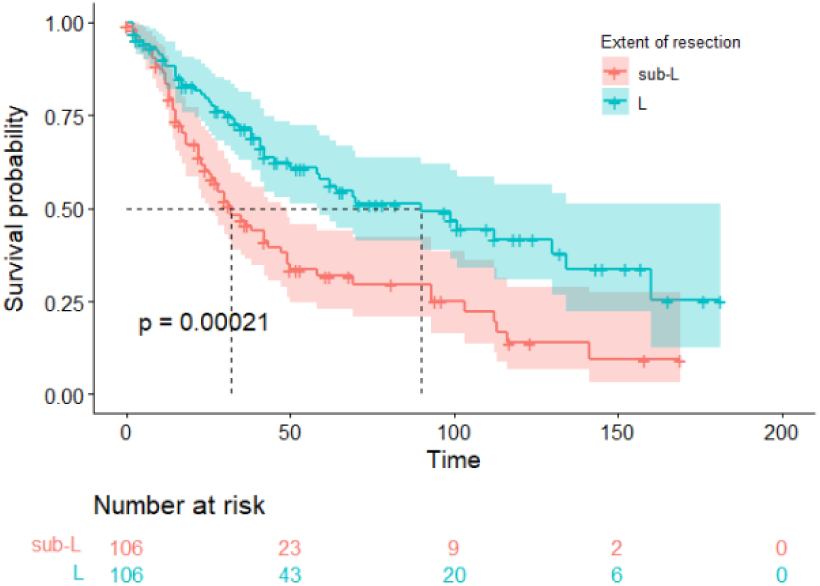
Kaplan–Meier survival curve of stage I SCLC patients with P-SCLC.

**Figure 6b.**
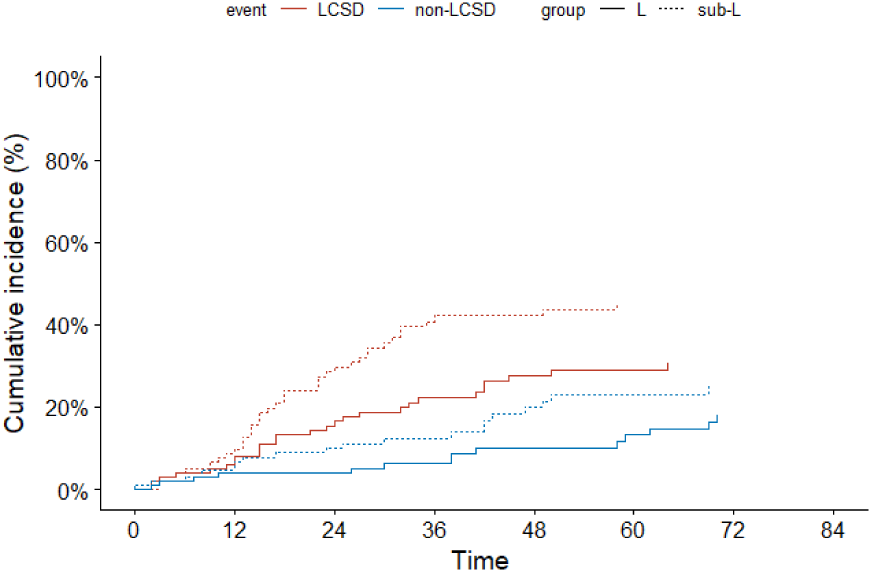
Cumulative incidence function curve of stage I SCLC patients with P-SCLC.

**Figure 6C.**
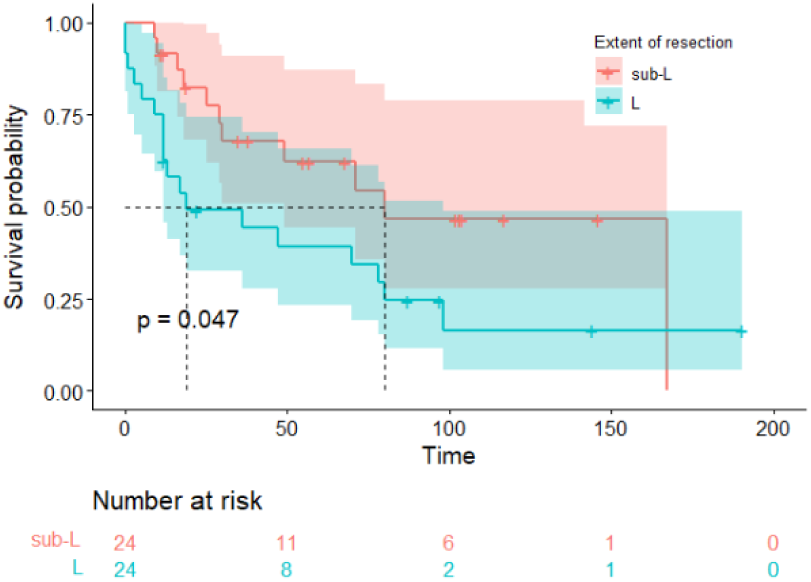
Kaplan–Meier survival curve of stage I SCLC patients with C-SCLC.

**Figure 6d.**
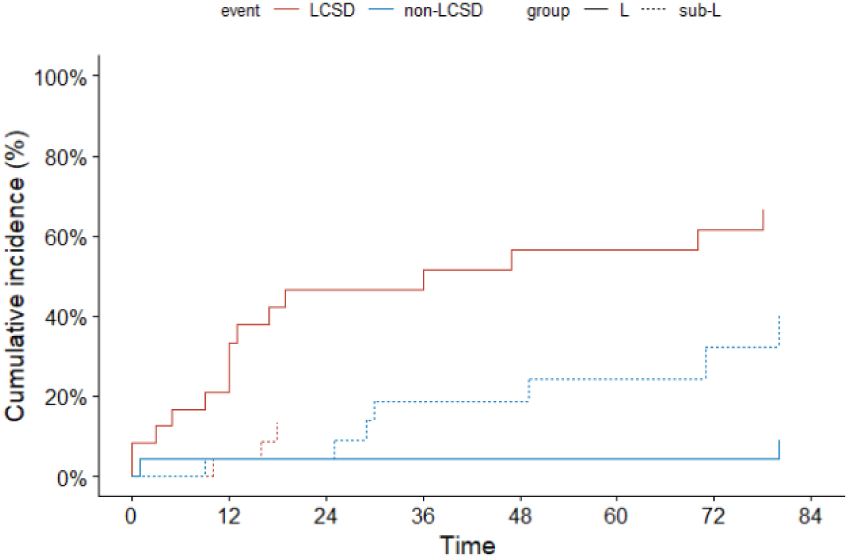
Cumulative incidence function curve of stage I SCLC patients with C-SCLC.

## Discusion

Multimodal therapy, including surgery, is increasingly embraced by thoracic surgeons for early-stage SCLC. The extent of resection remains controversial for early-stage SCLC due to limited evidence. Several studies utilizing the SEER database have explored influence of extent of resection on prognosis in limited SCLC [21–24]. Weksler et al. focused on stage I and II SCLC patients and found that those undergoing lobectomy or pneumonectomy had significantly longer survival than those opting for wedge resection. Similarly, Liu and colleagues reported improved survival with lobectomy compared to sublobar resection in T1-2N0M0 SCLC patients. However, Gu et al. analyzing pT1N0M0 SCLC patients, concluded that lobectomy showed similar overall and cancer-specific survival to sublobar resection. This corroborates the results of Zhou et al., whose study involving 188 stage I SCLC patients reached a similar conclusion. Two studies used NCBD database to examine outcomes of surgical approach in SCLC [25,26]. Combs et al. utilizing the NCDB, found that sublobar resection was associated with worse adjusted OS compared to lobar resection in patients with stage I-IIIA SCLC. In an analysis of cT1-2N0M0 SCLC patients using the NCDB database, Raman et al. distinguished wedge resection (WR) and segmental resection (SR) as separate entities. They found lobectomy exhibited similar survival to SR and superior survival to WR.

We observed a limited number of studies incorporating PSM in the past, which has constrained the significance of the conclusions drawn. Hence, this investigation seeks to leverage PSM to mitigate selection bias and provide a more accurate exploration of the potential role of surgical type in early-stage SCLC. Sublobar resection is typically performed in patients with poor physical condition or significant comorbidities, particularly respiratory diseases, potentially resulting in an increased likelihood of non-LCSD which further influencing OS outcome that past studies often exclusively concentrated on. Unfortunately, the SEER database lacks representation of this physical status-related baseline characteristic, posing a challenge for PSM in eliminating this bias. Although the lung-cancer specific survival (LCSS) was used to overcome this deficiency in two studies, they included competing events as censored data, which may have caused some bias [21,22]. In response, our study introduces competing risk model analysis for the first time, aiming to discern differences in LCSD between the L and sub-L groups.

CALGB140503 and JCOG0802, conducted as three-phase randomized controlled trials with a higher level of evidence, were the first to demonstrate the non-inferiority of sublobar resection compared to lobectomy in NSCLC. The main difference between CALGB140503 and JCOG0802 is the choice of sublobar resection technique. Although both techniques are allowed in CALGB140503, JCOG0802 requires the use of segmental resection. Despite producing consistent results, segmental resection is generally considered to be oncologically superior due to its ability to separate parenchyma along the intersegmental plane, allowing for a more thorough assessment of margins and lymph nodes [27]. In contrast, wedge resection is valued for its convenience, simplicity, and preservation of greater lung tissue. Firstly, we consider segmental resection and wedge resection as a whole, just like in the CALGB140503 study. Our findings indicate that lobectomy is associated with significantly improved OS and similar LCSD compared to sublobar resection in stage I SCLC patients. When differentiating between wedge resection and segmental resection as distinct procedures, it was noted that the overall survival of wedge resection was similar to that of segmental resection but inferior to lobectomy, which contrasts with Raman et al’s findings. After conducting competing risk model analysis, all three surgical procedures exhibited comparable LCSD. Our findings demonstrate the oncological equivalence of sublobar resection and offer the prospect of surgical treatment for patients with poor lung function or severe comorbidities.

Most SCLC are P-SCLC, but some may combine with additional components of any histological types of NSCLC, referred to as C-SCLC. Previous studies usually treated P-SCLC and C-SCLC as a whole. In our investigation, we explored the impact of the extent of resection on survival outcomes in P-SCLC and C-SCLC separately. In our analysis, we found no significant difference in overall survival (OS) and lung cancer-specific death (LCSD) outcomes between C-SCLC and P-SCLC. Several previous studies have demonstrated the impact of extent of resection on survival prognosis differs between C-SCLC and P-SCLC [27–30]. Our research shows that lobectomy was associated with increased OS and decreased LCSD compared to sublobar resection. While, to our surprise, the survival prognosis of lobectomy was significantly worse than sublobar resection in C-SCLC. The reason would be:(1) the low incidence of operation especially sublobar resection of C-SCLC results in a small sample size and low statistical power; (2) interpatient heterogeneity due to the additional components of any histological types of NSCLC.

The limitations of the study are as follows. Firstly, although we applied the PSM method to reduce selection bias as much as possible, we could only balance the available variables but not potentially unknown factors that may have affected the results. Secondly, despite SEER database is a population-based data, many clinicopathological variables are unavailable, such as lung function, comorbidities, adequacy of resection margin and recurrence-free survival (RFS), which play important roles in survival. Thirdly, the analysis was significantly constrained by the small size of the SR cohort, leading to a type I error. Finally: the SEER database lacked information on clinical stage, which can effectively mitigate staging bias arising from the uncertainty of lymph node dissection during sublobar resection.

## 5. Conclusion

In summary, our findings revealed that sublober resection is equal to lobectomy in lung cancer specific prognosis for the stage I patients. Sublober resection was suitable for the specifically selected patients with SCLC. But these findings still need to be verified by further prospective researches.

## Author contribution Funding

The authors received no specific funding for this work.

## Data availability declaration

The datasets used and/or analysed during the current study available from the corresponding author on reasonable request.

